# Prevalence and Clinical Determinants of Cognitive Impairment in Diverse Patients with Parkinson Disease

**DOI:** 10.64898/2026.06.02.26354673

**Authors:** Alexandra Zirra, Kamalesh C Dey, Ellen Camboe, Essa Bhadra, Rhiannon Laban, Brook Huxford, Shafaq Hussain-Ali, Cristina Simonet, Caroline Budu, David A Gallagher, Sheena Waters, Thomas Boyle, Viktoria Azoidou, Andrew J Lees, Maria Teresa Periñan, Charles R Marshall, Alastair J Noyce

## Abstract

**Importance:** The real-world prevalence and the clinical determinants associated with cognitive impairment in diverse patients with Parkinson disease (PD) have been understudied.

**Objective:** To determine the prevalence of cognitive impairment in a diverse PD cohort and explore associations with vascular, motor, and nonmotor factors.

**Design, setting and participants:** Case-only analysis of diverse patients with PD recruited to the East London Parkinson disease project (July 2022 to July 2025) at the Royal London Hospital, a tertiary referral center. Of 237 patients with cognitive status defined by expert, multi-disciplinary, clinical consensus, 223 remained after excluding atypical or secondary parkinsonism, other dementias, and study withdrawal.

**Exposures:** Observational study (no experimental intervention); exposures included vascular risk factors, motor and nonmotor clinical features.

**Main Outcome(s) and Measure(s):** The main outcome was cognitive impairment (PDCI), defined as mild cognitive impairment (PDMCI) or dementia (PDD) by expert clinical consensus based on clinical, imaging, and cognitive screening.

**Results:** Among 223 participants with a median disease duration of 4.0 (1.0-9.0) years, 112 (50.2%) had PDCI, including 62 (27.8%) with PDD and 50 (22.4%) with PDMCI. South Asian ethnicity was associated with PDCI in univariate analysis (OR, 2.30; 95% CI, 1.32-4.00, *P* = .003) and the association strengthened after adjusting for age, gender, years of education, disease duration and depression scores (OR, 3.60; 95% CI, 1.68-7.69, *P* < .001). PDCI was associated with increased odds of smoking (OR, 3.62; 95% CI, 1.56-8.41, *P* = .003) in the adjusted model. Increased odds were also associated with motor severity (Movement Disorders Society Unified Parkinson Disease Rating Scale Part III; OR per point increase 1.07; 95% CI, 1.04-1.10; *P* < .001), and daytime somnolence score (Epworth Sleepiness Scale; OR per point increase, 1.08; 95% CI, 1.01-1.16; *P* = .03).

**Conclusions and Relevance:** In this multi-ethnic study of PD using gold-standard expert multidisciplinary consensus, cognitive impairment was common and more prevalent among South Asian individuals. Smoking, greater motor severity, and higher daytime somnolence were associated with increased odds of cognitive impairment.

## Introduction

Cognitive impairment is one of the most feared complications of Parkinson disease (PD), leading to reduced quality of life and high caregiver burden.^1,2^ The spectrum of cognitive impairment ranges from subjective cognitive decline to mild cognitive impairment and dementia.^3^ Parkinson disease dementia (PDD) is characterized by a significant decline in more than one cognitive domain severe enough to impair activities of daily living.^4^ Mild cognitive impairment in PD is characterized by a decline in one or more cognitive domains but without a significant impact on the activities of daily living.^5^

The estimated prevalence of cognitive impairment in PD varies widely. Cross-sectional studies suggest a prevalence of cognitive impairment of approximately 50%, including both PDD and mild cognitive impairment.^3^ However, a large prospective international study, PPMI, reported a much higher frequency of 61.8% at 1 year, increasing to 69.9% at 4 years of follow up.^6^ Other studies, conducted in predominantly White, higher-income populations, have reported a prevalence of dementia of 17% at 5 years and mild cognitive impairment in 40-50% at 5 years from diagnosis.^3^ Notably, few studies included ethnically or socioeconomically diverse populations, limiting generalizability.^7^ Many of these studies used MDS Taskforce Level I diagnostic criteria, which are based primarily on recommended screening scales,^8^ rather than Level II comprehensive neuropsychological assessments.^5,9^ This reliance on screening based criteria may contribute to misclassification or overestimation of cognitive impairment in PD. This is particularly important in sociodemographically diverse groups in whom structural inequities and cultural or educational differences may bias screening outcomes independent of underlying neuropathology.

The recommended screening instruments for level I diagnosis of PDD and mild cognitive impairment include the Montreal Cognitive Assessment scale (MoCA), the Parkinson Disease-Cognitive Rating Scale and the Mattis Dementia Rating Scale Second Edition.^8^ The MoCA is the most widely used cognitive screening tool in dementia research, and increasingly used in PD.^10^ However, the original cut-off score of 26 for cognitive impairment, proposed in 2005, is now questioned in real-world clinical settings due to language-, cultural- and education-related biases.^8,11,12,13^

Several clinical factors have been associated with cognitive impairment in PD.^14,15,16^ Modifiable risk factors that overlap with the Lancet Commission for Dementia prevention include vascular risk factors, depression, motor severity, and hearing loss.^17^ Non-modifiable factors include older age, male sex, and duration of disease.^3^ Other studies have also identified minority ethnic background, lower levels of education, and socio-economic status as important factors associated with PDD based on Level I criteria.^18,19^

The East London Parkinson disease (ELPD) project aims to provide a robust real-world perspective of cognitive impairment in PD by using an expert multidisciplinary (MDT) consensus approach. We examined vascular, motor, nonmotor and social factors associated with cognitive impairment in a multiethnic, socioeconomically diverse cohort, using consecutive recruitment, deep phenotyping, standardized clinical assessment and transparent analytic methods, in a setting where conventional screening instruments for cognitive impairment may be suboptimal. We hypothesized that the prevalence of cognitive impairment would be higher in ethnically and socioeconomically diverse patients with PD and could be driven by vascular risk factors.

## Methods

### Study Design and Participants

The East London Parkinson disease project is a case-control study, approved in November 2018 (REC: 39/W/0255),^20^ conducted in a single tertiary centre setting. Patients with PD and parkinsonism from the Movement Disorder outpatient clinic at the Royal London Hospital were recruited consecutively. The present analysis represents a cross-sectional, case-only substudy conducted between July 2022 and July 2025, focusing on in-depth cognitive screening for patients with PD.

The inclusion criteria were: patients over the age of 18, with a clinical diagnosis of PD by experienced movement disorders specialists, able to consent or with an appropriate proxy for consent. To maximize inclusivity across cultures and severity of cognitive impairment, the ELPD project offered home visits and permitted proxy consent for those with diminished capacity due to cognitive impairment. Exclusion criteria were: atypical parkinsonism, secondary parkinsonism, other dementia (not attributable to neurodegenerative or cerebrovascular causes) and withdrawal of consent to participate.

### Data Collection

Two coupled clinical visits were undertaken for all participants. As described in our previous publication,^20^ visit 1 focused on demographic and clinical characteristics of PD, including motor and nonmotor assessments, and a brief cognitive screening with the MoCA. Visit 2 focused on in-depth cognitive screening tests, including the Clinical Dementia Rating scale (CDR).^21^ Ethnicity was self-reported as per 2021 UK Census categories^22^ and grouped as White, South Asian, or Other.

Socioeconomic status was assessed using the Index of Multiple Deprivation, a national area-based measure of deprivation, and expressed as national deciles. Decile 1 represents the most deprived, and decile 10 the least deprived.^23,24^

### Outcomes and Exposures

The main outcome for this study was cognitive impairment defined by MDT consensus. The multidisciplinary team consisted of movement disorder neurologists, a cognitive neurologist, a clinical nurse specialist and the clinical researchers conducting the assessments. For each case, we took into consideration: demographic information - including native language, years of education, ethnicity, and age, clinical characteristics - duration of disease, cognitive or neuropsychiatric complaints from clinical letters or from researchers’ interaction, comorbidities and current medication, and investigations - MoCA, CDR, depression and anxiety scores (Hospital Anxiety and Depression scale), and, where available, neuroimaging (114/150 [76%]). Each case obtained an outcome of PD normal cognition (PDNC), PD mild cognitive impairment (PDMCI), PD dementia (PDD) or other - eFigure 1 in Supplement. The main analyses compared PDNC and PDCI (PD cognitive impairment - consisting of PDD and PDMCI patients). Secondary analyses comparing PDNC and PDD were performed. A total of 223 patients were retained, out of 237 patients screened (eFigure 2 in Supplement provides details of exclusions).

The primary exposures included ethnicity and vascular risk factors: hypercholesterolemia, hypertension, type 2 diabetes and smoking status. Secondary exposures included motor severity (Movement Disorders Society Unified Parkinson Disease Rating Scale Part III, MDS-UPDRS Part III), and non-motor features: daytime somnolence (Epworth Sleepiness Scale, ESS), sleep disturbance (Parkinson Sleep Scale version 2, PSS2), clinically reported probable REM-sleep behaviour disorder (RBD) status, probable RBD score (REM sleep Behaviour Disorder Screening Questionnaire, RBDSQ), olfaction (6-item abbreviated smell test), hearing loss, and socioeconomic deprivation (Index of Multiple Deprivation decile, IMD)^24–29^.

### Statistical Analysis

Statistical analysis and figure generation were performed in Jupyter Lab v 3.4.3 using Python v 3.13. Data were imputed using single imputation (median for continuous variables and mode for binary variables) when missingness was <5%, assuming missing at random. Data were not imputed when missingness was >5%. For the descriptive tables, Chi-squared tests were used for categorical variables and Mann Whitney U tests for continuous variables (eTable 1 in Supplement). Sample size calculations for prevalence used Epitools Epidemiological Calculators assuming PDCI prevalence of 45% (White) and 73% (South Asian) based on previous MoCA data; with α=0.05, 90% power, and 1:1 allocation, 70 participants per group were required.^20,30^ Univariate logistic regression models with cognitive status as the outcome were undertaken for covariate exploration. Odds ratios (ORs) with 95% confidence intervals (CIs) were reported. All tests were 2-sided, and statistical significance was defined as *P* <.05. Predicted probabilities were calculated and plotted for continuous covariates from fitted logistic regression models.

For primary and secondary exposures, univariate logistic regression models were run initially. Multivariable logistic regression models were adjusted for the following covariates: age, gender, ethnicity, years of education, disease duration and depression scores (Hospital Anxiety and Depression Scale-Depression subscale [HADS-D] - Depression). For the South Asian ethnicity exposure, the same covariates were used except for ethnicity. Covariates were chosen after evaluating collinearity, including only those with a variance inflation factor (VIF) below 2 and a Pearson correlation coefficient (ρ) below 0.5. The linearity of the logit was evaluated for continuous covariates and exposures, with all variables satisfying the assumption (*P* > .05). Due to high missingness, for specific exposures (RBDSQ, PSS2, 6-item smell test, and hearing loss) a reduced multivariable model with the same covariates was explored.

Pseudoanonymised data were stored as ‘.csv’ files, securely on Queen Mary University of London servers are available through the Verily Workbench (GP2 tier 2 access). Analysis code will be made publicly available on GitHub: https://github.com/Wolfson-PNU-QMUL/ELPD_cognitive_impairment_prevalence.

## Results

### Prevalence of cognitive impairment in a diverse population

From 237 patients with available cognitive outcomes, 223 patients were retained for the analysis with a median (IQR) age of 70.0 (62.5-76) years, and median (IQR) disease duration 4.0 (1.0-9.0) years. Men numbered 127/223 (57.0%) and ethnicity was diverse - 98/223 (43.9%) White, 86/223 (38.6%) South Asian, and 39/223 (17.5%) Other. The Other category included 17 Black (12 African, 5 Caribbean), 11 Middle Eastern, 7 Asian and 4 mixed ethnicity patients.

The median (IQR) age in patients with PDCI was 74.0 (67.8-80.0) years, which was older than 66.0 (58.0-73.0) years for PDNC patients. Patients with PDCI had fewer years of education and higher depression scores compared with those with PDNC (Table 1).

**Table 1.** Demographics in the two groups: PDNC - Parkinson Disease Normal Cognition, PDCI - Parkinson Disease Cognitive Impairment. Continuous variables expressed as median (IQR), categorical variables as n (%). LEDD - Levodopa Equivalent Daily Dose, Hchol – hypercholesterolemia, HTN – hypertension, T2DM – type 2 diabetes mellitus. RBDSQ – REM sleep behaviour screening questionnaire, PSS2 – Parkinson sleep scale version 2, IMD - Index of Multiple Deprivation, RBDSQ - REM sleep Behaviour Screening Questionnaire, PSS2 – Parkinson Sleep Scale version 2. a, Mann-Whitney U test, b, Chi-Squared. * Driven by White vs South Asian

| Variables | PDNC<br>n=111 | PDCI<br>n=112 | PDCI vs PDNC |
| --- | --- | --- | --- |
| Age, years | 66.0 (58.0-73.0) | 74.0 (67.8-80.0) | <.001<sup>a</sup> |
| Gender |  |  |  |
| Female | 42/111 (37.8) | 54/112 (48.2) | .153<sup>b</sup> |
| Male | 69/111 (62.2) | 58/112 (51.8) |  |
| Ethnicity |  |  |  |
| White | 62/111 (55.9) | 36/112 (32.1) | .001<sup>b*</sup> |
| South Asian | 32/111 (28.8) | 54/112 (48.2) |  |
| Other | 17/111 (15.3) | 22/112 (19.6) |  |
| Years of education | 13.0 (11.0-16.0) | 10.0 (7.8-12.0) | <.001<sup>a</sup> |
| Time since diagnosis, years | 3.0 (1.0-8.0) | 5.0 (1.0-9.2) | .214<sup>a</sup> |
| Time from symptom onset, years | 6.0 (3.0-11.0) | 7.0 (4.0-12.0) | .231<sup>a</sup> |
| LEDD, mg | 600.0 (300.0-800.0) | 600.0 (300.0-831.2) | .541<sup>a</sup> |
| HADS Depression scores | 3.0 (2.0-7.0) | 8.0 (5.0-11.0) | <.001<sup>a</sup> |
| Smoking status (ever smokers) | 25/109 (22.9) | 39/109 (35.8) | .053<sup>b</sup> |
| Hchol | 42/111 (37.8) | 70/112 (62.5) | <.001<sup>b</sup> |
| T2DM | 21/111 (18.9) | 39/112 (34.8) | .011<sup>b</sup> |
| HTN | 47/111 (42.3) | 67/112 (59.8) | .013<sup>b</sup> |
| MDS-UPDRS Part III | 23.0 (16.0-30.0) | 47.0 (32.0-61.0) | <.001<sup>a</sup> |
| ESS score | 6.0 (2.0-9.0) | 8.5 (5.0-13.0) | <.001<sup>a</sup> |
| RBD status | 20/110 (18.2) | 31 (27.7) | .128<sup>b</sup> |
| IMD | 3.0 (2.0-5.0) | 3.0 (2.0-4.0) | .004<sup>a</sup> |
| RBDSQ score | 2.5 (1.0-6.0) | 4.0 (1.0-7.0) | .101<sup>a</sup> |
| PSS2 score | 12.0 (7.0-23.0) | 16.0 (10.0-27.0) | .010<sup>a</sup> |
| Hearing loss | 21/104 (20.2) | 34/98 (34.7) | .031<sup>b</sup> |
| Smell test | 3.0 (2.0-4.0) | 2.0 (1.0-3.0) | <.001<sup>a</sup> |
| South Asian ethnicity | 32/86 (37.2) | 54/86 (62.8) | .005<sup>b</sup> |

The cross-sectional prevalence of PDNC was 111/223 (49.8%) and of PDCI 112/223 (50.2%), with PDD diagnosed in 62/223 (27.8%). Among South Asian participants, 54/86 (62.8%) were classified as having PDCI and 32/86 (37.2%) as having PDNC.

### Univariate associations with PDCI

In univariate logistic models, South Asian ethnicity (OR, 2.30; 95% CI, 1.32-4.00; *P* = .003), higher depression (HADS-D, OR per point increase, 1.25; 95% CI, 1.16-1.35; *P* < .001), higher age (OR per year increase 1.10; 95% CI, 1.06-1.13; *P* < .001) and fewer years of education (OR per year increase, 0.86, 95% CI, 0.80-0.92, P<.001) were associated with increased odds of PDCI (eFigure 3A in Supplement). The index of multiple deprivation decile was weakly inversely associated with PDCI (OR per point increase, 0.88; 95% CI, 0.79-0.99; *P* = .029).

Hypercholesterolemia, type 2 diabetes, hypertension and smoking were all associated with increased odds of PDCI (eFigure 5A in Supplement). Greater motor severity (MDS-UPDRS Part III: OR per point increase, 1.08; 95% CI, 1.06-1.11; *P* < .001) and non-motor features, particularly higher daytime somnolence (ESS: OR per point increase, 1.12; 95% CI, 1.06-1.18; *P* < .001), were also associated with PDCI. Associations with sleep quality and hearing loss were weaker, while RBD features were not significantly associated. Lower smell test scores were strongly associated with increased odds of PDCI (OR per point increase, 0.70; 95% CI, 0.58-0.85; *P* < .001).

### Multivariable associations with PDCI

South Asian ethnicity was independently associated with PDCI (OR 3.60; 95% CI, 1.68-7.69; *P* < .001) after adjustment for age, gender, education, disease duration and depression (Figure 1). This association remained robust after additional adjustment for vascular risk factors, smoking, motor severity, non-motor features and IMD (Figure 2).

**Figure 1.**
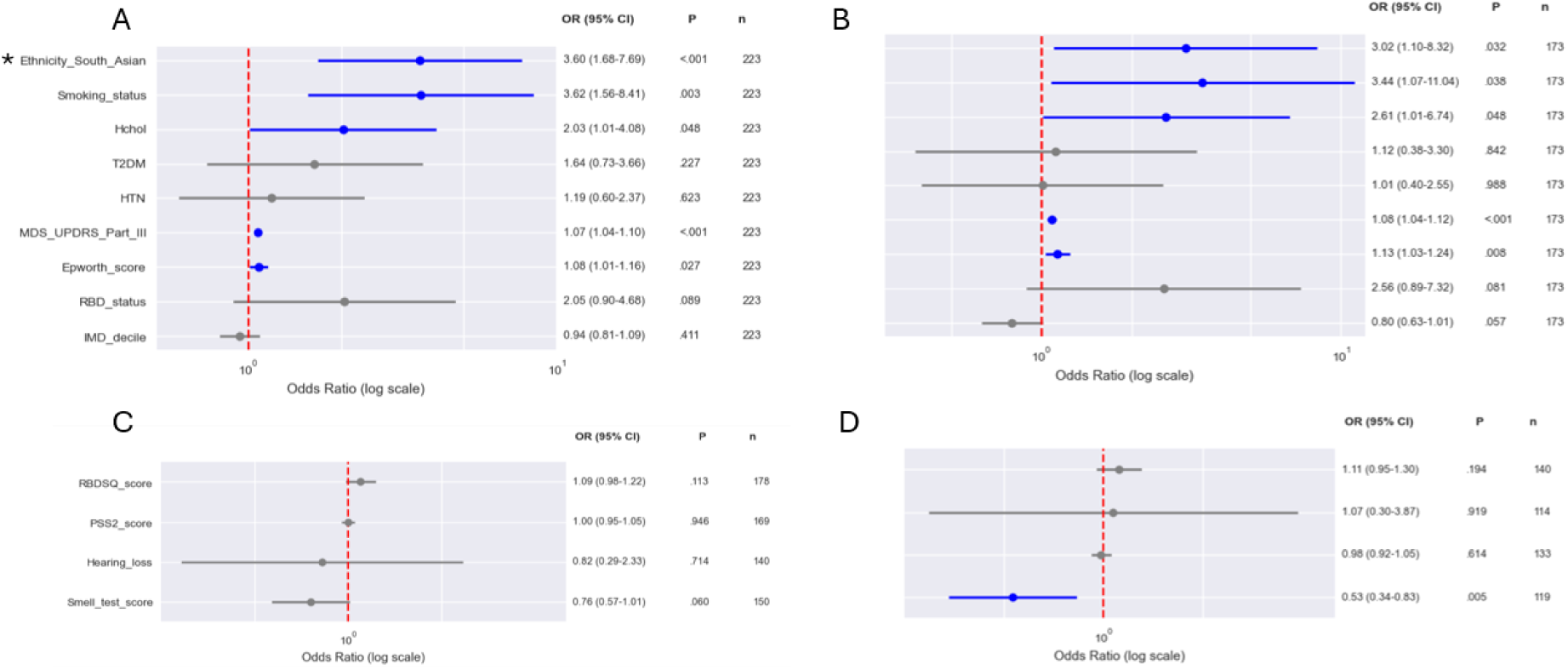
Multivariable logistic regression for primary and secondary exposures in the PDCI full model (A) and reduced (C), PDD full model (B) and reduced (D). Model adjusted for age, gender, ethnicity, education years, duration of disease, and depression scores. * - model not adjusted for ethnicity. Hchol –hypercholesterolemia, HTN – hypertension, T2DM – type 2 diabetes mellitus. RBDSQ – REM sleep behaviour screening questionnaire, PSS2 – Parkinson sleep scale version 2, OR - odds ratio, 95% CI - 95% confidence interval. Colour shading indicated statistical significance (P<.05)

**Figure 2.**
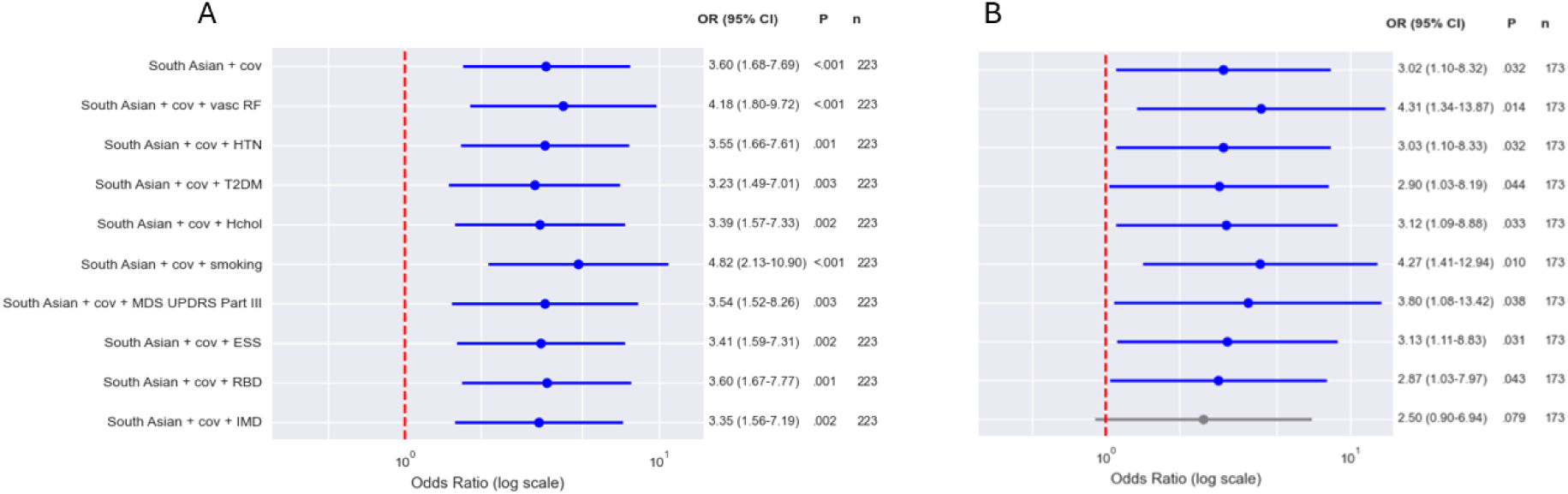
Multivariable logistic regression for South Asian exposure in the PDCI model (A) and PDD model (B). cov – covariates, vasc RF – vascular risk factors, HTN – hypertension, T2DM – type 2 diabetes mellitus, Hchol – hypercholesterolemia, ESS – Epworth Sleep Scale, RBD – REM sleep behaviour disorder, IMD – index of multiple deprivation decile, OR - odds ratio, 95% CI - 95% confidence interval. Color shading indicates statistical significance (P < .05).

Among vascular factors, smoking (OR 3.62; 95% CI, 1.56-8.41; *P* = .003) and hypercholesterolemia (OR 2.03; 95% CI, 1.01–4.08; P = .048) were independently associated with PDCI, whereas hypertension and type 2 diabetes were not (Figure 1A).

Greater motor severity (MDS-UPDRS Part III: OR per point increase, 1.07; 95% CI, 1.04-1.10; *P* < .001) and worse daytime somnolence (ESS: OR per point increase, 1.08; 95% CI, 1.01-1.16; *P* = .027) were also associated with PDCI. Other factors, including sleep measures, IMD and hearing loss, were not associated after adjustment (Figure 2).

### Univariate and multivariable associations with PDD sub-analyses

In univariate analyses, PDD was associated with older age, South Asian ethnicity, higher depression scores, fewer years of education, and longer disease duration (eFigure 3B, eTable 2 in Supplement). Greater motor severity, daytime somnolence, RBD features, lower smell test scores, hearing loss and IMD were also associated.

In multivariable analyses, South Asian ethnicity remained associated with PDD (OR 3.02; 95% CI, 1.10-8.32; *P* = .032), but this was attenuated after adjustment for IMD (OR per point increase 2.50; 95% CI, 0.90-6.94; *P* = .079) (Figure 2B).

Overall, patterns were similar to PDCI, with associations observed for vascular risk factors, motor severity and daytime somnolence, while lower smell test scores were specifically associated with PDD.

## Discussion

In this multiethnic, socioeconomically diverse East London Parkinson disease cohort, PDCI affected over half of participants and PDD was present in over a quarter of participants. Both outcomes were associated with approximately threefold higher odds in South Asian patients, with the prevalence of cognitive impairment approximately 15% more prevalent than in White participants. These associations were not attenuated after adjustment for vascular risk factors. Cognitive impairment was linked to older age, lower education, higher depressive symptoms, while disease duration was specifically associated with dementia in univariate models. Smoking, motor severity and excessive daytime somnolence were associated with both outcomes, while lower olfaction scores were linked only to dementia. Hypercholesterolemia showed a weak association with both.

Previous ELPD data reported PDCI prevalence estimates of 73% in South Asian and 75% in Black participants, compared with 45% in White participants, using standard MoCA testing.^20^ Using a robust expert consensus approach, our study found a smaller but strongly significant difference in South Asian participants (62.8%) compared with White patients (36.7%), suggesting our approach mitigated to some extent the cultural bias of pen and paper cognitive screening tools.

Prevalence of PDCI at 4 years was comparable to UK community studies, suggesting our cohort is representative of the underlying population structure.^31^ However, a higher prevalence might have been anticipated given the tertiary care setting, which typically includes patients with more complex disease, as well as greater ethnic and socioeconomic diversity. Cognitive impairment has been reported as higher in ethnically diverse participants, such as in Black and Latino or Hispanic patients.^32,33^ However, accurate ascertainment is particularly challenging in these underrepresented groups. Brief screening tools, such as the MoCA, identify global cognitive deficits but may overestimate impairment due to language, cultural and educational differences.^34,35^ In comparison, MDS Task Force Level II criteria require comprehensive neuropsychological assessment for greater diagnostic specificity.^36^ Although mitigation strategies for screening tools exist, they are not universally accessible.

South Asian ethnicity was significantly associated with cognitive impairment in both the PDCI and PDD models. Biological mechanisms may contribute to this, including vascular risk factors and apolipoprotein E (*APOE*) carrier status, which has been linked to dementia and dyslipidaemia across diverse populations.^37,38,39^ *APOE*-related mechanisms may partly explain the higher PDCI prevalence observed, although this remains speculative. The association persisted after adjustment for vascular, motor, and nonmotor factors. For hypercholestorolemia, the analyses excluded lipid fractions, likely obscuring the granularity of vascular exposure. In contrast, adjustment for socioeconomic deprivation attenuated the association between South Asian ethnicity and PDD, suggesting that socioeconomic rather than biological factors may contribute to cognitive disparities in underrepresented UK populations. This is consistent with evidence that ethnicity and area-level deprivation are independently associated with all-cause dementia risk in diverse UK populations, suggesting that highlighting the need for targeted assessment in underrepresented populations.^40^

We also investigated vascular risk factors in isolation and found a strong association between smoking history and PDCI, which conflicts with the established protective association of smoking with PD risk.^41,42^ Type 2 diabetes and hypertension were not associated with PDCI, despite their high prevalence and prior literature suggesting their contribution to cognitive impairment in PD.^43,44,14^ These null findings should be interpreted cautiously given cohort heterogeneity, differences in age and socioeconomic characteristics, and sample size limitations.

Our findings are consistent with previous reports identifying increased odds of cognitive impairment in PD with older age, longer disease duration, greater motor severity, and more severe daytime somnolence.^3^ In contrast, we found no significant association with RBD status. Previous studies using polysomnography-confirmed RBD have reported links with cognitive decline.^45,46,47^ The use of questionnaire-based probable RBD diagnoses after PD onset in our work may have introduced misclassification and attenuated associations. Alternatively, non-Lewy body pathologies, such as cerebrovascular disease, may be substantial drivers of cognitive impairment in this population, explaining the attenuated association with RBD.

Strengths of this study include robust real-world characterization of cognitive impairment using expert consensus and comprehensive clinical phenotyping, improving diagnostic specificity beyond brief screening tools. This approach mitigated potential biases by accounting for patients’ native language, education, age, comorbidities, medication burden, and multiple screening tests. Translated scales were used where possible, and available imaging helped exclude other causes of dementia.

Another strength of our study was recruitment from diverse backgrounds. Barriers to inclusion of underrepresented populations include poor awareness and misconceptions about research, as well as disease-related difficulties^48^ which we addressed through an inclusive framework involving community engagement and bilingual researchers (Dey KC et al, unpublished data).

One limitation of this study is recruitment from a single tertiary centre, which may limit generalizability, particularly to rural populations. However, our cohort was diverse in terms of ethnicity, socioeconomic background, and education, which partially mitigates this concern. Secondly, the cross-sectional design limits causal inference and may introduce reverse causation for non-motor and vascular factors. Conversely, prospective studies often suffer from healthy participant bias, which we mitigated through our robust recruitment strategies. Thirdly, using binary categories for some exposures and missing data for others may have introduced sampling biases. Finally, potential model overfitting, particularly for PDD models with a limited number of events, represents an additional limitation, which is why PDD analyses were conducted as secondary rather than primary analyses.

## Conclusion

Using an expert multidisciplinary consensus approach, we demonstrated a high prevalence of cognitive impairment at a median of 4 years from diagnosis, with higher rates among South Asian participants. These differences may reflect a complex interplay of cultural, socioeconomic, and educational factors rather than ethnicity alone, though causal inference cannot be drawn from this cross-sectional analysis. Cognitive impairment was associated with older age, greater motor severity, and smoking, while other vascular risk factors showed no independent links, possibly reflecting residual confounding. These findings highlight the need for culturally sensitive, methodologically rigorous assessment of cognition in PD, particularly in underrepresented populations. Prospective and retrospective longitudinal studies are needed to clarify temporal and causal relationships.

## Acknowledgments

We thank all participants and their families for contributing to this study. We are grateful to KCD for substantial support with participant recruitment, and to AJN, CM, MTP and VA for extensive first-stage review and feedback on the manuscript. We also acknowledge the clinical and research staff at Barts Health NHS Trust and affiliated community partners, as well as our laboratory team for assistance with sample processing and data management.

## Contributors

AJN, AZ and CM conceived and designed the study. AZ performed data analysis and wrote the first draft. KCD, EB, EC and AZ contributed substantially to participant recruitment. AJN, CM, MTP and VA conducted the initial review of data and manuscript drafts. All authors contributed to data interpretation, critically revised the manuscript, and approved the final version.

## Access to data and data analysis

Only the named study team members (AJN, AZ, KCD, EC, EB) had full access to the study data. AZ performed the data analysis under supervision of AJN, CM and MTP.

## Data sharing statement

Available through the Verily Workbench, Global Parkinson’s Genetics Program (GP2) - tier 2 access. GP2 is funded by the Aligning Science Across Parkinson’s (ASAP) initiative and implemented by The Michael J. Fox Foundation for Parkinson’s Research (https://gp2.org). All GP2 data for these analyses is available through collaboration with Accelerating Medicines Partnership in Parkinson’s disease and is available through application on the AMP-PD website (https://amp-pd.org/register-for-amp-pd).

## Supplementary

**eTable 1.** PD missingness for covariates and exposures. N total – total number of participants, N missing – total number of missing values for each variable, Pct missing – total percentage of missingness for each variable, PDCI - Parkinson Disease Cognitive Impairment, HTN - Hypertension, T2DM - Type 2 Diabetes Mellitus, Hchol - Hypercholesterolemia, RBD - REM-sleep Behaviour Disorder, ESS - Epworth Sleepiness Score, MDS-UPDRS - Movement Disorders Society United Parkinson Disease Rating Scale, HADS - Hospital Anxiety and Depression Scale, PSS2 - Parkinson Sleep Scale version 2.

| Variable | Type | Min | Max | N total | N missing | Pct missing |
| --- | --- | --- | --- | --- | --- | --- |
| Age | Continuous | 35.0 | 92.0 | 223 | 0 | 0.0 |
| Ethnicity | Categorical | 1.0 | 3.0 | 223 | 0 | 0.0 |
| Gender | Categorical | 0.0 | 1.0 | 223 | 0 | 0.0 |
| Education years | Continuous | 0.0 | 24.0 | 223 | 0 | 0.0 |
| Disease duration | Continuous | 0.0 | 33.0 | 223 | 0 | 0.0 |
| PDCI | Categorical | 0.0 | 1.0 | 223 | 0 | 0.0 |
| HTN | Categorical | 0.0 | 1.0 | 223 | 0 | 0.0 |
| T2DM | Categorical | 0.0 | 1.0 | 223 | 0 | 0.0 |
| Hchol | Categorical | 0.0 | 1.0 | 223 | 0 | 0.0 |
| RBD status | Categorical | 0.0 | 1.0 | 223 | 1 | 0.4 |
| ESS score | Continuous | 0.0 | 24.0 | 223 | 2 | 0.9 |
| MDS-UPDRS Part III | Continuous | 3.0 | 109.0 | 223 | 5 | 2.2 |
| Smoking status | Categorical | 0.0 | 1.0 | 223 | 5 | 2.2 |
| HADS Depression score | Continuous | 0.0 | 21.0 | 223 | 6 | 2.7 |
| Hearing loss | Categorical | 0.0 | 1.0 | 223 | 21 | 9.4 |
| Smell test score | Continuous | 0.0 | 6.0 | 223 | 23 | 10.3 |
| RBDSQ score | Continuous | 0.0 | 13.0 | 223 | 45 | 20.2 |
| PSS2 score | Continuous | 0.0 | 40.0 | 223 | 53 | 23.8 |

**eTable 2.** Demographics of the two groups: PDNC - Parkinson disease with normal cognition, PDD - Parkinson disease dementia. Continuous variables expressed as median (IQR), categorical variables as n / n total (%). a, Mann-Whitney U, b, Chi-Squared. LEDD - Levodopa Equivalent Daily Dose, HADS - Hospital Anxiety and Depression Scale, Hchol - Hypercholesterolemia, T2DM - Type 2 Diabetes Mellitus, HTN - Hypertension, MDS-UPDRS - Movement Disorders Society United Parkinson Disease Rating Scale, ESS - Epworth Sleepiness Sclae, RBD - REM-sleep Behaviour Disorder, IMD - Index of Multiple Deprivation, RBDSQ - REM-sleep Behaviour Disorder Screening Questionnaire, PSS2 - Parkinson Sleep Scale version 2.

| Variables | PDNC<br>n=111 | PDD<br>n=62 | PDD vs<br>PDNC |
| --- | --- | --- | --- |
| Age, years | 66.0 (58.0-73.0) | 76.0 (72.0-81.8) | <.001<sup>a</sup> |
| **Gender** |  |  |  |
| **Female** | 42/111 (37.8) | 32/62 (51.6) | .111<sup>b</sup> |
| **Male** | 69/111 (62.2) | 30/62 (48.4) |  |
| **Ethnicity** |  |  |  |
| **White** | 62/111 (55.9) | 22/62 (35.5) | .028<sup>b</sup> |
| **South Asian** | 32/111 (28.8) | 29/62 (46.8) |  |
| **Other** | 17/111 (15.3) | 11/62 (17.7) |  |
| **Years of education** | 13.0 (11.0-16.0) | 10.0 (5.0-12.0) | <.001<sup>a</sup> |
| **Time since diagnosis, years** | 3.0 (1.0-8.0) | 6.0 (1.2-11.8) | .031<sup>a</sup> |
| **Time from symptom onset, years** | 6.0 (3.0-11.0) | 8.0 (5.0-13.0) | .027<sup>a</sup> |
| **LEDD, mg** | 600.0 (300.0-800.0) | 600.0 (375.0-843.8) | .395<sup>a</sup> |
| **HADS Depression scores** | 3.0 (2.0-7.0) | 9.0 (6.0-12.0) | <.001<sup>a</sup> |
| **Smoking status**<br>**(ever smokers)** | 25/109 (22.9) | 20/62 (32.3) | .250<sup>b</sup> |
| **Hchol** | 42/111 (37.8) | 41/62 (66.1) | <.001<sup>b</sup> |
| **T2DM** | 21/111 (18.9) | 20/62 (32.3) | .073<sup>b</sup> |
| **HTN** | 47/111 (42.3) | 37/62 (59.7) | .042<sup>b</sup> |
| **MDS-UPDRS Part III** | 23.0 (16.0-30.0) | 53.5 (37.0-71.2) | <.001<sup>a</sup> |
| **ESS score** | 6.0 (2.0-9.0) | 11.0 (6.0-14.0) | <.001<sup>a</sup> |
| **RBD status** | 20/110 (18.2) | 20/62 (32.3) | .056<sup>b</sup> |
| **IMD** | 3.0 (2.0-5.0) | 3.0 (2.0-3.0) | <.001<sup>a</sup> |
| **RBDSQ score** | 2.5 (1.0-6.0) | 5.0 (1.0-7.0) | .070<sup>a</sup> |
| **PSS2 score** | 12.0 (7.0-23.0) | 17.0 (10.8-26.2) | .018<sup>a</sup> |
| **Hearing loss** | 21/104 (20.2) | 25/58 (43.1) | .003<sup>b</sup> |
| **Smell test** | 3.0 (2.0-4.0) | 1.0 (0.0-2.0) | <.001<sup>a</sup> |
| **South Asian ethnicity** | 32/86 (37.2) | 29/86 (33.7) | .028<sup>b</sup> |

**eFigure 1.**
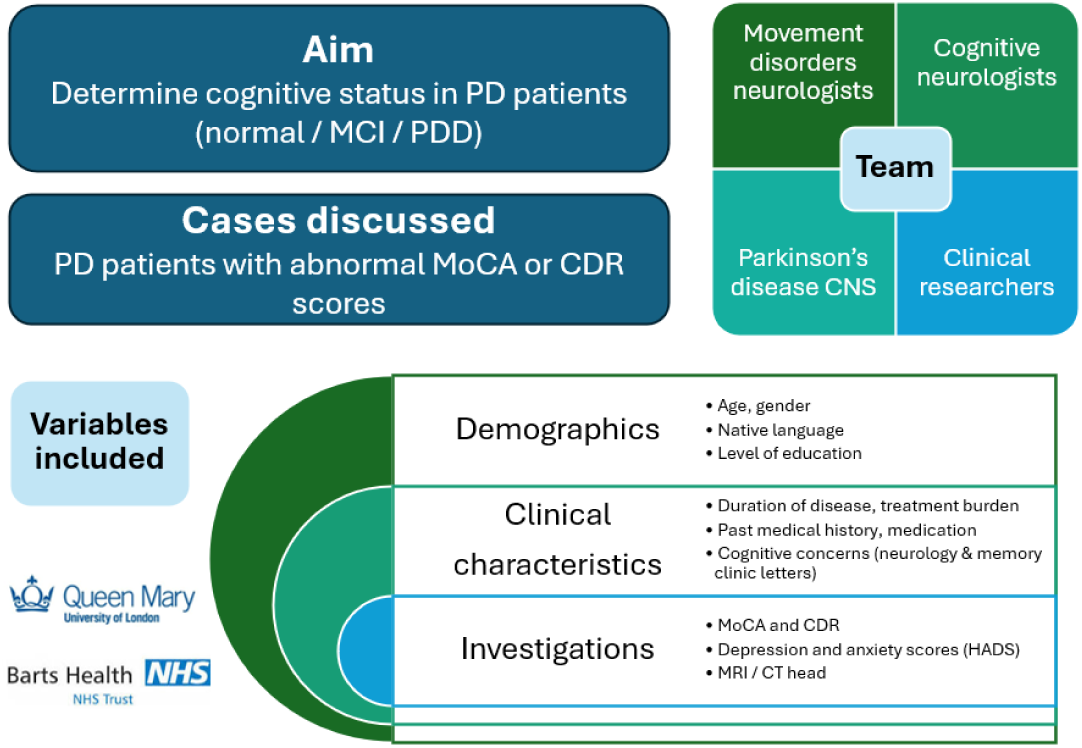
Expert multidisciplinary consensus meeting structure for establishing cognitive outcomes.

**eFigure 2.**
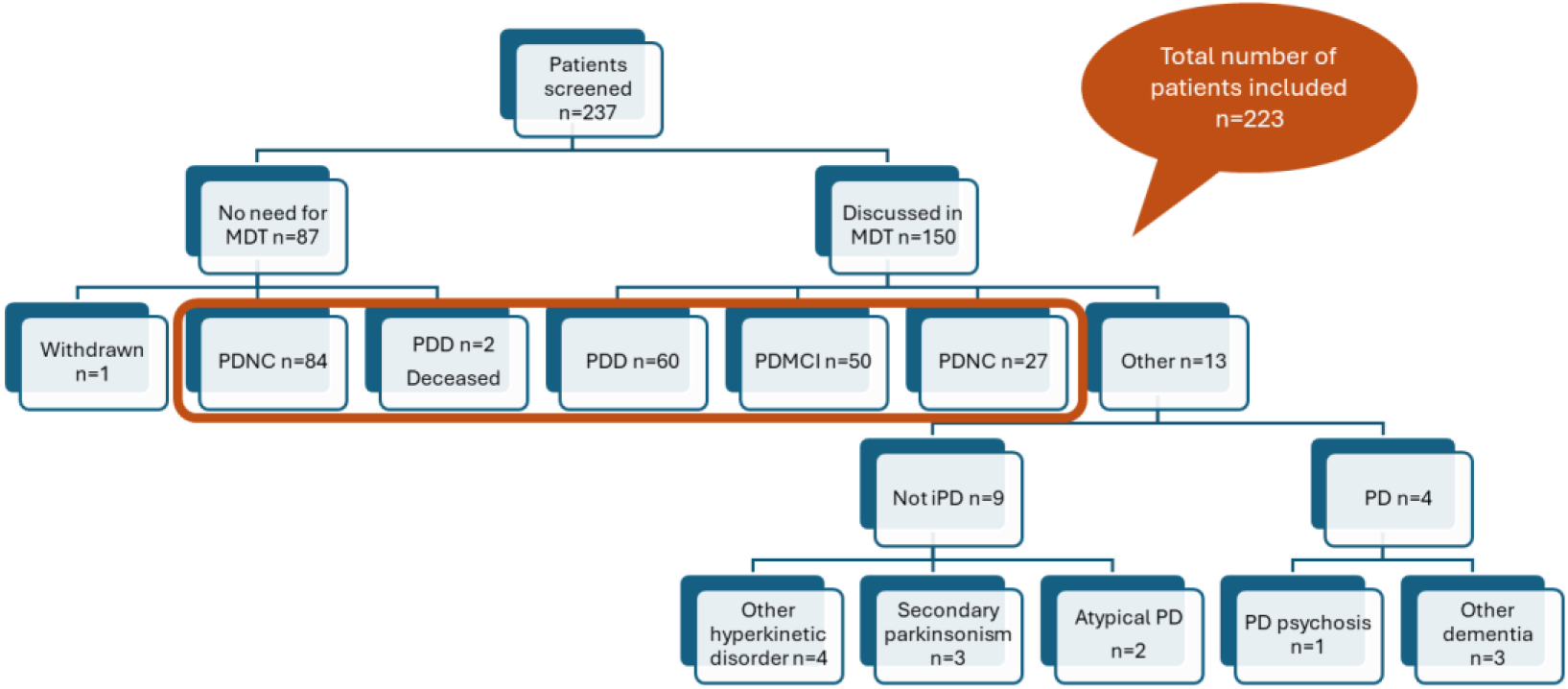
Expert consensus outcomes. PD – Parkinson Disease, PDD – Parkinson Disease Dementia, PDMCI – Parkinson Disease Mild Cognitive Impairment, PDNC – Parkinson Disease Normal Cognition, MDT – expert multidisciplinary consensus meeting.

**eFigure 3.**
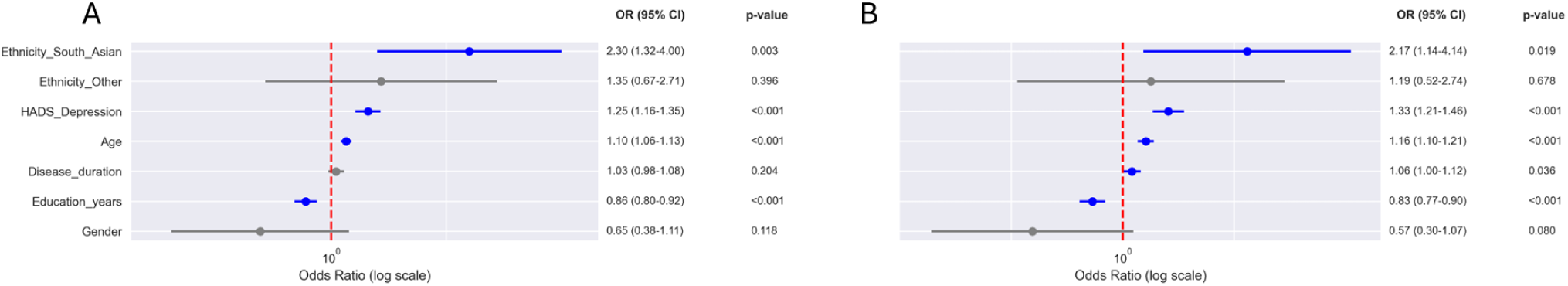
Univariate logistic regressions for covariates in (A) PDCI (n=223) and (B) PDD (n=173). Color shading indicates statistical significance (P < .05). HADS - Hospital Anxiety and Depression Scale.

**eFigure 4.**
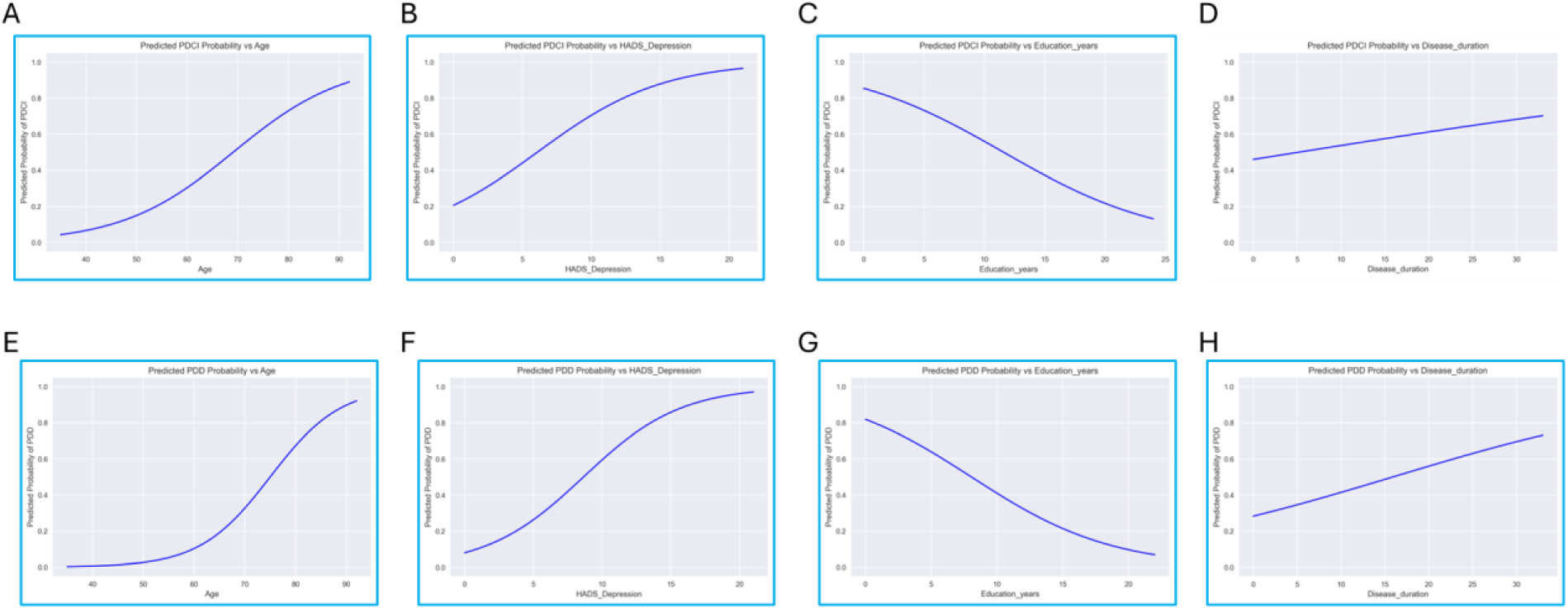
Predicted PDCI probability (A-D) and PDD probability (E-H) versus age, HADS depression score, years of education, and disease duration. Colour shading indicated statistical significance (P<.05) in univariate models. PDCI - Parkinson Disease Cognitive Impairment, PDD - Parkinson Disease Dementia, HADS - Hospital Anxiety and Depression Scale.

**eFigure 5.**
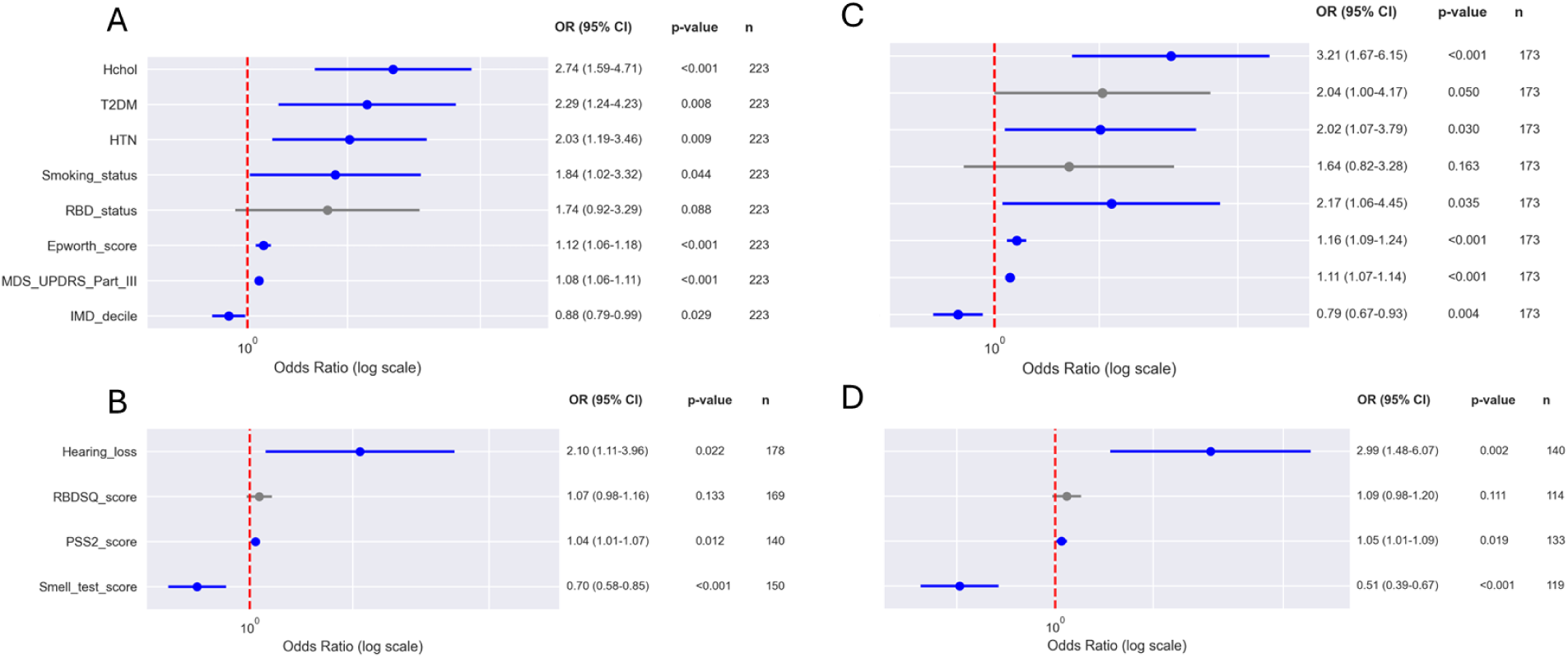
Univariate logistic regressions for exposures in the PDCI full model (A) and reduced model (B), PDD full model (C) and reduced model (D). Color shading indicates statistical significance (P < .05). PDCI - Parkinson Disease Cognitive Impairment, PDD - Parkinson Disease Dementia, Hchol - Hypercholesterolemia, T2DM - Type 2 Diabetes Mellitus, HTN - Hypertension, RBD - REM-sleep Behaviour Disorder, ESS - Epworth Sleepiness Scale, MDS-UPDRS - Movement Disorders Society United Parkinson Disease Rating Scale, IMD - Index of Multiple Deprivation, RBDSQ - REM-sleep Behaviour Disorder Screening Questionnaire, PSS2 - Parkinson Sleep Scale version 2.

